# Delta/ Omicron and BA.1/BA.2 co-infections occurring in Immunocompromised hosts

**DOI:** 10.1101/2022.04.04.22273058

**Authors:** Richard Leuking, Madhusudhanan Narasimhan, Lenin Mahimainathan, Alagar R Muthukumar, Yan Liu, Chao Xing, Christian P. Larsen, Andrew Clark, Jeffrey A. SoRelle

**Affiliations:** Department of Internal Medicine, University of Texas Southwestern Medical Center, Division of Infectious Diseases and Geographic Medicine, Dallas, TX, USA; Department of Pathology, University of Texas Southwestern Medical Center, Dallas, Texas, USA; McDermott Center for Human Growth and Development, University of Texas Southwestern Medical Center, Dallas, Texas

## Abstract

Concomitant infection of multiple SARS-CoV-2 variants has become an increasing concern, as this scenario increases the likelihood of recombinant variants. Detecting co-infection of SARS-CoV-2 variants is difficult to detect by whole genome sequencing approaches, but genotyping methods facilitate detection. We describe 2 cases of Delta/Omicron and 2 cases of Omicron sublineage BA.1/ BA.2 co-infection as detected by a multiplex genotyping fragment analysis method. Findings were confirmed by whole genome sequencing. Review of the patient characteristics revealed co-morbidities and conditions which weaken the immune system and may make them more susceptible to harboring SARS-CoV-2 variant co-infections.

## Introduction

Ongoing genomic surveillance has revealed the trajectory of severe acute respiratory syndrome coronavirus 2 (SARS-CoV-2) infections largely coincide with the emergence of dominant genetic lineages (variants) harboring distinct genetic mutations. SARS-CoV-2 variants of concern (VOCs) have arisen, harboring a high number of mutations and increased infectivity, morbidity, immune evasion, and mortality among infected individuals. Evolutionarily, the SARS-CoV-2 virus mutates relatively slowly compared to HIV or influenza RNA viruses owing to the presence of nonstructural protein 14 (Nsp14) that harbors 3’-to-5’ exoribonuclease (ExoN), a unique RNA proof-reader not found in other RNA viruses. Genome-wide association studies (GWAS) indeed indicate that mutations in error-correcting nsp14 protein exhibited a strongest association with increased SARS-CoV-2 genome-wide mutation load (1-3). Other factors can also change the rate of evolution as VOCs rise to high frequencies.

For instance, when multiple variants circulate, the possibility of genomic recombination between different strains or sub-lineages increases. Recombination potentially occurs when two distinct lineages are present in the same host. High levels of recombination may occur sporadically in RNA viruses and recombination may not be a trait optimized by natural selection. Importantly, these novel genotypes are concerning and may result in a more virulent virus in a short amount of time. Indeed, this was recently observed to occur when the Omicron variant out-competed the Delta VOC (4). In addition, we have observed that severely immunocompromised (IC) hosts with no antibody immune response may harbor the virus for up to a year with no increased mutation rate. However, some IC hosts may exhibit an immune response low enough to prevent viral clearance, but that is high enough select for viral variation. Such a scenario has been observed in uncontrolled HIV patients with low but detectable CD4 counts or other transplant patients (5-7).

While a combination of genetic drift, recombination, and population immunity could play a key role in the continual emergence of VOCs, reasons underlying this emergence remain poorly understood. In the face of population immunity, we and others have previously reported that in IC patients, viruses elude vaccines or remain more pathogenic (8-10). Further, Sun et al.

(11) have shown that IC individuals exhibit a higher risk of COVID-19 breakthrough infection. Of note, weakened immune system was determined to be a potential source for multiple amino acid substitutions and deletions in the spike protein, with increased resistance to neutralizing antibodies (5). Thus, these studies suggest IC states prolong viral survival and provide opportunities for additional mutations. It is thus critical to address whether underlying immunodeficiency or immunosuppressive therapy impacts the emergence of recombinant variants by permitting co-infection of variants.

Typically, molecular tools such as whole genome sequencing (WGS) have facilitated evolutionary and epidemiological monitoring of SARS-CoV-2 in near real-time and enabled us to better prepare and combat the threat. Detection and characterization of co-infection and emerging recombinant variants by current WGS methods is challenging with respect to obtaining and analyzing complete viral genomes from clinical samples in a timely and cost-effective manner. However, genotyping methods allow sensitive detection of multiple mutations simultaneously, which can provide a clue to the existence of co-infection. Using fragment analysis genotyping method for detecting SARS-CoV-2 variants (12), we detected two cases of a Delta/Omicron co-infection and two cases of an BA.1/BA.2 co-infection. These findings were confirmed by WGS.

The present manuscript presents two cases of SARS-CoV-2 Delta/Omicron (B.1.617.2/ B.1.1.529) co-infection, and the first two cases of co-infection with BA.1/BA.2 Omicron sub-lineages among IC patients. The knowledge of co-infections is critical and will enforce an evidence-based understanding of the mechanisms driving genomic variability, patterns of emergence, and the clinical management and treatment of COVID-19 cases with co-infection.

### Case Description and Clinical Presentation of the lung transplant patient

A male in his 70’s with an lung transplant presented with fever and respiratory symptoms in November 2021 (Figure 1). SARS-CoV-2 PCR was positive and SARS-CoV-2 genotyping identified mutations consistent with the Delta variant. The patient’s outpatient immunosuppression regimen included prednisone, tacrolimus, and monthly belatacept infusions. He had received three doses of the Pfizer/BioNTech COVID-19 vaccine prior to presentation. The patient was initially treated with casirivimab/ imdevimab (Regeneron) followed by dexamethasone and remdesivir for 5 and 10 days, respectively. He was discharged without supplemental oxygen but reported continued exertional dyspnea and reduced forced expiratory volume (FEV1) from baseline.

**Figure 1.**
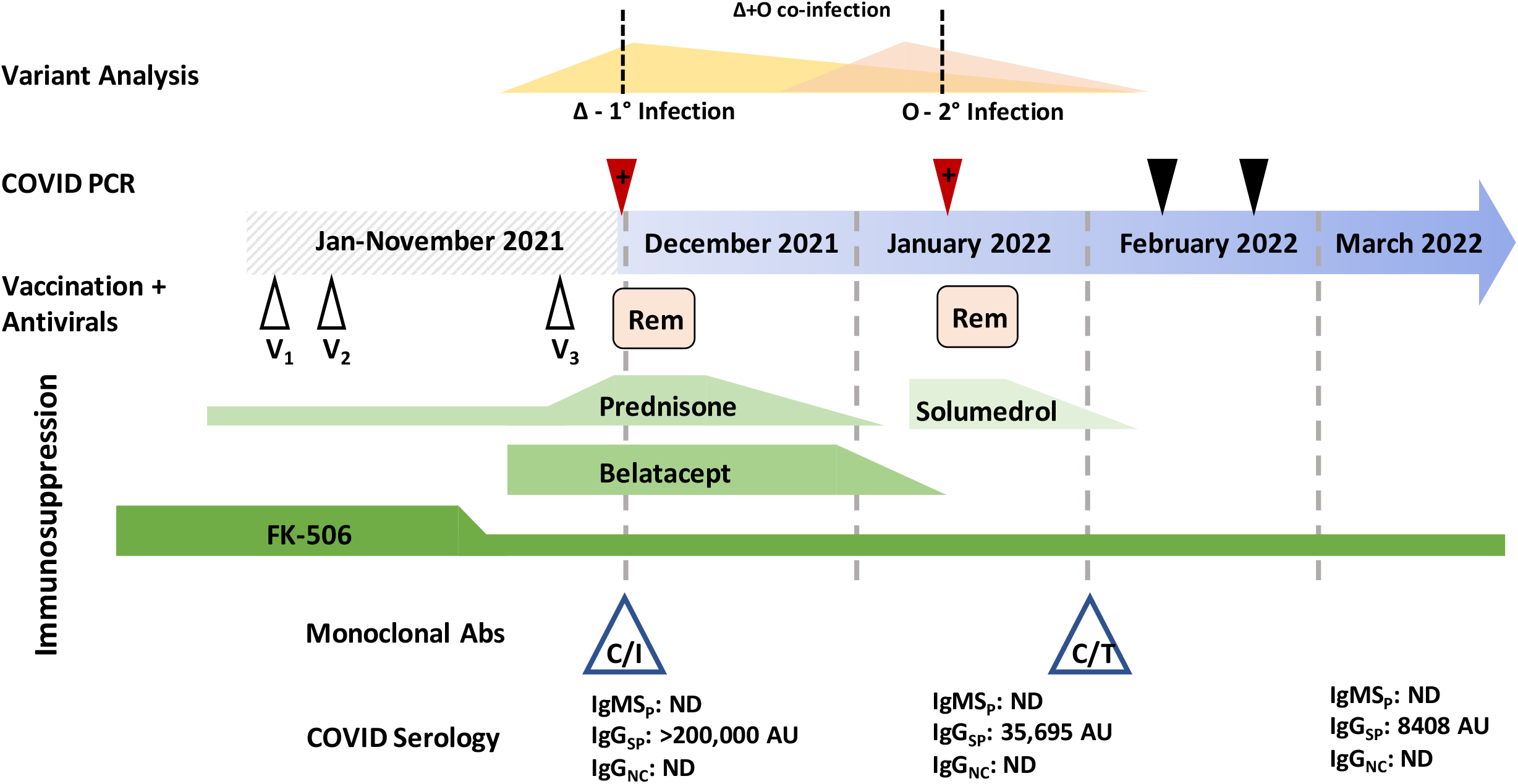
Variant analysis was performed at the points marked by a red triangle (positive SARS-CoV-2 test) and dashed line above intersects the hypothesized viral loads of a Delta (Δ) and Omicron (O) variant in each sample. Black triangles are negative SARS-CoV-2 PCR tests. V_1_-V_3_ represent vaccine doses. Rem: remdesivir. Green lines indicate immunosuppressants and the height of the shape reflects the dose level. Monoclonal antibody treatment and administration time is indicated by the blue triangles. C/I: casirivimab/imdevimab, C/T: tixagevimab and cilgavimab. COVID serology indicates when specific antibody tests were performed. IgM_SP_: IgM anti-spike, IgG_SP_: IgG anti-spike, IgG_NC_: IgG anti-nucleocapsid, ND: not detected, AU: arbitrary units.

Six weeks later (early January 2022), he returned with fever, worsening shortness of breath, and productive cough. The case patient again showed SARS-CoV-2 PCR positivity, but given the proximity to prior infection, was initially thought to be a result of persistent viral shedding. The patient was treated for hospital acquired pneumonia with piperacillin-tazobactam and vancomycin and methylprednisolone for acute cellular rejection. However, SARS-CoV-2 genotyping of the second PCR-positive specimen identified the presence of two SARS-CoV-2 variants: Delta (remaining from his initial infection) and Omicron (suggestive of a secondary infection) (Figure 2A). As the Omicron variant had not yet emerged when the patient had initial SARS-CoV-2 infection, the genotyping and epidemiologic data indicated a novel infection on top of a persistent Delta infection. With confirmation of active SARS-CoV-2 disease, a course of dexamethasone and remdesivir was recommended to improve the symptomology in the case patient. The patient was also given tixagevimab and cilgavimab treatment for further prophylaxis. With discontinued antibiotics and tapered steroids, the patient was discharged without supplemental oxygen and a FEV1 near baseline.

**Figure 2.**
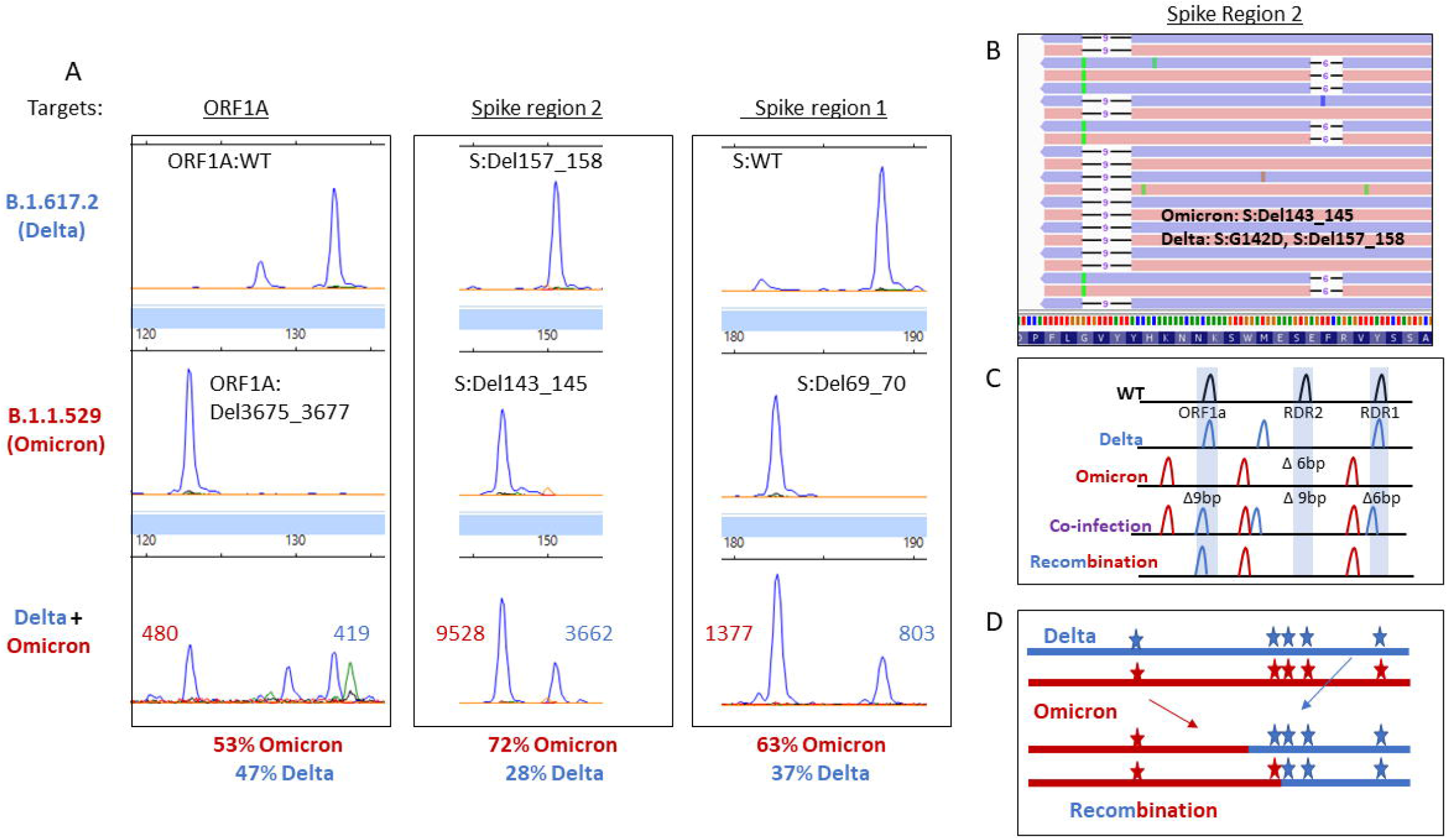
A) Electrophoretogram of the ORF1A, Spike region 1 and Spike region 2 sites where different sized amplicons indicative of Delta (top) Omicron (middle) or co-infection (bottom) are present. B) Next-generation sequencing reads were visualized in the Integrated Genome Viewer with read direction colored red or blue. Deletions are indicated by a bar, single nucleotide variants have the variant nucleotide present, and insertions are highlighted by a purple box. C) Schematic illustrating the expected location of fragments for variants under normal, mixed or recombined conditions.

### Defective Nucleocapsid IgG response despite infection

During both infections, the patient had undetectable IgG and IgM antibodies against nucleocapsid (Figure 1), which is useful for distinguishing infection-associated immune response from vaccine-related response (13). He was given monoclonal antibody therapy (casirivimab and imdevimab) during both infections, resulting in very high Spike IgG antibody levels. Notably, casirivimab and imdevimab are human monoclonal antibodies that bind to nonoverlapping epitopes of the S protein RBD of SARS-CoV-2 (anti-SARS-CoV-2 antibodies). The lack of nucleocapsid antibody response reflects an inability to mount an antibody response due to immunosuppression. Several studies have previously reported that patients with pre-existing immunosuppressive conditions or those who undergo immunosuppressive treatments exhibit low or no ability to mount antibody response (8,9).

### Additional Cases of SARS-CoV-2 Co-infections

Following the identification of the primary case reported in this work, we continued surveillance of all positive SARS-CoV-2 specimens for additional cases of SARS-CoV-2 co-infections with different VOCs. While we report the above case in the greatest level of clinical detail, we have also encountered another specimen from a female in her 70’s with uncontrolled diabetes had Delta/Omicron coinfection (diagnosed 1st week of January 2022, had received 3 mRNA vaccinations). Only a single sample was available but returned the same results when re-extracted to rule out contamination.

We next identified three independent patients with BA.1/ BA.2 co-infection (Table 1, collected last week of February and last week of March). One was a woman in her 60’s with a hematologic malignancy after 5 rounds of chemotherapy and had no COVID-19 vaccinations. She initially tested positive for COVID-19 in early January 2021 at an outside institution when BA.1 was the predominant variant and was treated with sotrovimab monoclonal antibody therapy. Her symptoms persisted and worsened in early March when the co-infection was identified. Another BA.1/BA.2 co-infection occurred in a man in his 50’s with all three mRNA COVID-19 vaccinations (most recent was a little over 90 days before his infection) but no significant medical history aside from a stroke. The last patient was a female in her 80’s who had chronic lymphocytic leukemia and was started on anti-B cell therapy (obinutuzumab and venetoclax) after her initial two doses of mRNA vaccine. She was still on these therapies when she received her mRNA booster in 6 months before this infection.

**Table 1.** Patient characteristics

| Age/ Sex | PMH | COVID Dx | Antibody testing | Treatment | Vaccination |
| --- | --- | --- | --- | --- | --- |
| 70's M | Lung transplant | November<br>January | NC IgG/ Sp IgM:<br>Not detected<br>November-March | November:<br>Casirivimab/imdevimab,<br>steroids, remdesivir<br>February: Tixagevimab<br>and cilgavimab | 3 doses mRNA |
| 70's F | Uncontrolled Diabetes type 2; HbA1C: 14 | January | None | Admitted 3 days for AKI.<br>No COVID treatments | 2 doses mRNA |
| 60's F | DLBCL s/p 5 cycles RCHOP (last cycle: Feb '21) | January<br>March | None | Sotrovimab, steroids (Jan), readmitted March (2 weeks) | No vaccination |
| 80's F | CLL on Venetoclax (Mar '21- Feb '22) and Obinutuzumab (Mar '21-Aug '21) | March | Spike IgG: 123 AU/mL (Oct '21) | Asymptomatic | 3 doses mRNA |
| 50's M | Asthma, stroke | March | None | Asymptomatic | 3 doses mRNA. |

Only a single sample from each of patient was available for testing, but re-extraction of each returned the same result revealed multiple peaks amplified for RDR1 (WT and 6b.p. deletion), RDR2 (WT and 9 b.p. deletion), RDR3-4 (WT and a 6 b.p. insertion) (Figure 3A). This pattern could occur with BA.1 and BA.2 variant genomic RNA present in the same reaction arising from either co-infection or contamination. Carry-over contamination has been a frequent issue in WGS methods but has not occurred in the fragment analysis PCR test (as determined during CLIA validation). To rule out the possibility of contamination, RNA was re-extracted from each sample and repeated. The confirmatory fragment analysis results were the same.

**Figure 3.**
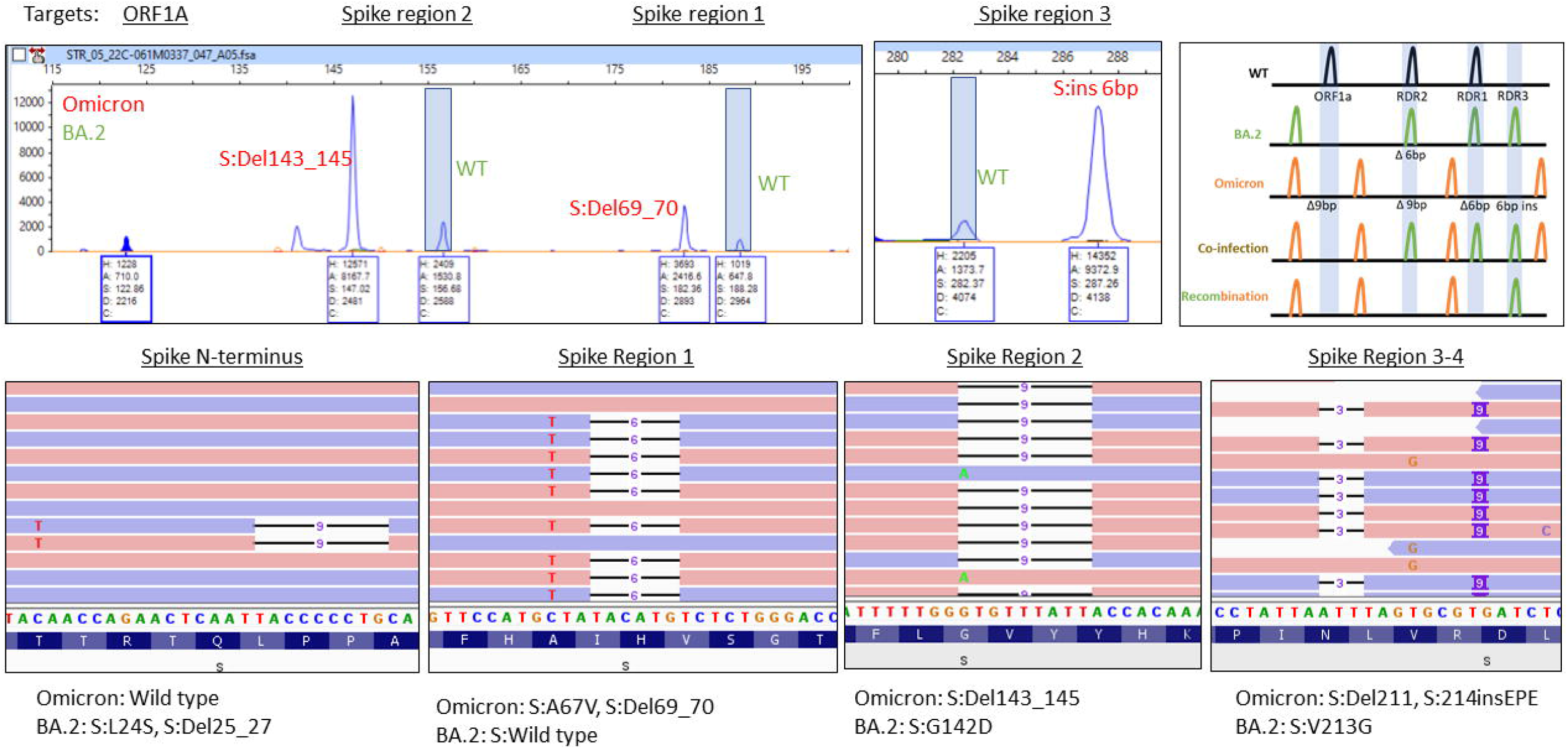
A) Electrophoretogram of ORF1A, Spike region 1, Spike region 2, and Spike region 3-4 sites where different sized amplicons indicative of Omicron (red) and BA.2 (green) co-infection are present. B) Schematic illustrating the expected location of fragments for variants under normal, mixed or recombined conditions. C) Next-generation sequencing reads were visualized in the Integrated Genome Viewer with read direction colored red or blue. Deletions are indicated by a bar, single nucleotide variants have the variant nucleotide present, and insertions are highlighted by a purple box.

The co-occurrence of amplicons specific to the BA.1 and BA.2 variants indicates a co-infection rather than recombination has occurred. Determining whether a co-infection is present by whole genome sequencing requires mutations be within 100-150 base pairs of each other to observe co-occurrence of mutations on the same read strand, we focused on the spike gene where multiple differences in mutations were found.

Sequencing showed separate forward and reverse reads (colored red and blue respectively, Figure 3B) for the BA.1mutations (S:69_70Del and A67V) and BA.2 (no mutations present) in the RDR1 region. In the RDR2 region, the BA.1 deletion (S:143_145Del) is present with alternative reads from the BA.2 variant (S:G142D). In the RDR3-4 region, separate reads are observed with mutations indicative of for BA.1 (S:211Del, 214 insEPE) and BA.2 (S:V213G). Lastly, in the N-terminal portion of the S gene, an S:L24S, S:25_27Del variants from BA.2 are present with sequence consistent with the BA.1 variant (no mutation).

### Laboratory findings and genetic relationship of the patient’s SARS-CoV-2 variants

Variant status is routinely determined for all SARS-CoV-2 PCR-positive samples at our institution through a combination of WGS and a genotyping PCR analyzed by fragment analysis. Briefly, for fragment analysis, mutational hotspots of the SARS-CoV-2 genome are amplified with fluorescently labeled primers. These PCR amplicons (fragments) are separated by capillary electrophoresis, and size differences determine the presence or absences of characteristic deletions. The initial PCR-positive specimen solely contained Delta variant sequences, while the patient’s subsequent specimen contained mutational signatures consistent with co-infection by the initial Delta VOC and the newly emerged Omicron VOC (B.1.1.529). Mutations specific to Delta and Omicron were found on independent reads using WGS (Figure 2B), which indicates non-recombined viral infection. While a total of eight fragment analysis targets are used, only three are highlighted to demonstrate the difference in Omicron and Delta variants. ORF1A detects the 3 amino acid deletion from ORF1A:Del3675_3677. Spike recurrently deleted region (RDR)1 and 2 detect S:Del69_70 and mutations in the 140-160 amino acid range, respectively. The Delta variant has a 2 amino acid deletion in Spike region 2 (S:Del157_158), while Omicron has a 3 amino acid deletion (S:Del143_145). No mutagenic signatures were identified by either genotyping or NGS to suggest recombination between the two lineages (Figure 2C).

## Discussion

Here we describe two cases of Delta/Omicron co-infection and two cases of BA.1/BA.2. While infectivity and replication dynamics of individual SARS-CoV-2 genetic lineages continue to be elucidated, significantly less is known concerning co-infections with unique SARS-CoV-2 variants. The paucity of information in the literature is likely multifactorial, given an epidemiological pattern of variant emergence and nonuniform application or access to sequencing or genotyping methods. Given the recent history of Delta-confirmed infection in November, the finding of Delta-Omicron co-infection appears authentic as a staggered type of co-infection and not the result of laboratory contamination. Testing of re-extracted sample helped rule-out laboratory contamination. In concordance with our findings, prior studies have also identified co-infection as an impetus for recombination between different SARS-CoV-2 variants (14,15).

As this is the first reported case report of BA.1/BA.2 variant co-infections so soon after an Omicron/Delta co-infection, we predict such co-infections will continue to occur when one variant takes over another. Specifically, this is likely to occur in IC hosts who have trouble clearing the virus, thus, allowing it to effectively adapt by exploiting the host cell conditions. In this context, host proteins, sirtuin 1 and 5 (SIRT1 and SIRT5) involved in energy, metabolism, and survival processes, have been recently shown to interact with SARS-CoV-2 Nsp14 and generating a lethal phenotype of SARS-CoV-2 with effective replication and/or long-term propagation (16). Importantly, the expression and activity of SIRT1 was shown to be enhanced by the steroid, prednisolone (17), which the case patient has been receiving since transplant at a dose of 7.5mg daily until the November COVID diagnosis and continued with 60 mg daily following this initial diagnosis. Thus, any such alterations in host cell pathways due to IC and/or IS drugs that can interfere with and dampen the SARS-CoV-2 nsp14-ExoN is likely to hamper the intrinsic fidelity and contribute to high-level mutagenesis.

The small number of cases observed prevents generalizable comments on whether co-infection protracts disease course or has more severe symptoms. The serological responses among IC patients are dampened as exemplified by the lack of Nucleocapsid IgG antibody response after months of primary and secondary infection in this lung transplant patient. The high anti-Spike IgG levels were due to monoclonal antibody use. The outcomes of these patients indicates unvaccinated and immunosuppressed patients are at higher risk for more severe disease.

A key learning point of these cases is that protracted disease presents an opportunity for additional exposure or the emergence of new variants. Of note, it has been reported that persistently infected IC patients accumulate amino acid substitutions or deletions in different regions of human SARS-CoV-2 spike protein and thus are conceived to be a source of new immune-escape viral variants (5,18,19). Also, CD4+ T-cell depletion (<20 CD4+ T-cell counts) as found in IC persons, including persons living with HIV/AIDS, has been determined to increase the risk of giving rise to SARS-CoV-2 resistance mutations (6-7). In contrast, an absent immune response does not reject virus and thus there is no competitive advantage for acquiring resistance mutations. Furthermore, we have learned that these cases of SARS-CoV-2 co-infection are most likely to occur in times when there is a diversity of variants in circulation. Thus, it is likely that co-infections will continue to occur and methods to monitor this should be considered.

The most commonly used variant detection method is currently WGS. However, criteria for calling variants are that mutations must be detected at >50% or higher levels of variant allele frequencies. Additionally, due to biases introduced in amplicon-based library preparation, whole genome sequencing cannot accurately quantify differences in variant levels. This cut-off prevents minor allele-frequencies to be counted for detection of co-infections. Genotyping methods offer a sensitive alternative where multiple mutations can be detected at once provided a wild-type and mutant allele can be detected simultaneously. In this scenario, fragment analysis is superior, because it can detect multiple types of deletions simultaneously. This method of multiplex SARS-CoV-2 genotyping by fragment analysis lends itself to not only detecting concurrent variant mutational signatures but also allows relative quantitation of the genomic material present. This quantitation can be cross applied to the CT value to infer whether an infection is active or not. For our patient, a 25% decrease in Delta nucleic acid corresponds with a 4-fold difference (2 CT value difference), which indicates the Delta variant was still active and not shedding dead virus.

Furthermore, genomic recombination can be screened by fragment analysis. The variant identification program, Pangolin, can distinguish recombination events if it has been trained to do, such as recently discovered recombinant variants named: XD, XE, and XF. Nextclade, non-amplicon based platforms and bioinformatic tools such as RAT (Recombination Analysis Tool) or RDP (Recombination Detection Program) may also help detect recombinant events. However, this fragment analysis assay has targets across the genome from ORF1A to spike and ORF8/ Nucleocapsid genes, where differences in a specific region may indicate recombination. Most assays just target the spike gene and recombination can occur across the entire SARS-CoV-2 genome.

Therefore, we conclude that SARS-CoV-2 co-infections will continue and represent a source of viral evolution, which in many cases can be effectively monitored using a fragment analysis genotyping approach.

## Data Availability

All data produced in the present work are contained in the manuscript

## Acknowledgements

The Texas SARS-CoV-2 Variant Network Project is funded and supported by the Texas Department of State Health Services (DSHS) as part of a financial assistance award from the Centers for Disease Control and Prevention (CDC) of the U.S. Department of Health and Human Services (HHS) totaling $15 million, with 100 percent funded by CDC/HHS. The contents are those of the author(s) and do not necessarily represent the official views of, nor an endorsement by, DSHS, CDC/HHS, or the U.S. Government.

